# The impact on mental health of young Asian Americans due to acts of discrimination and hate crimes during COVID-19

**DOI:** 10.1101/2023.01.13.23284501

**Authors:** Ahmed Gawash, David F. Lo, Brianna Nghiem, Priscilla Rofail, Sayan Basu

## Abstract

This article is an examination of the acts of discriminiation and hate crimes against the Asian American community and how their mental health has been impacted. The historical examination of acts against the Asian American community stem from acts against the Asian American community during both the yellow peril and the Roosevelt Era. Alongside current day institutionalized policies that are propagated by the media, the resurrection of historical tropes further act to seclude Asian Americans from mainstream society. These acts of seclusion further drive mental health inequality in Asian American society that include, but are not limited to: anxiety, depression, and psychological stress. These mental health inequalities are further subdivided into different ethnic groups that worsen as data is collected from older generations. More recently, COVID-19 has brought forth an upsurge in hate crimes that has only worsened the ability of Asian American youth to fully develop a racial identity; the upsurge in hate crimes is also coupled with invalidation of interethnic differences and invalidation of their discriminatory experiences. The synthesis of current day research will aid in the understanding of the mental health inequality in the Asian American community and aid in further studies that can address these plights.

**Purpose Statement:** The purpose of this paper is to investigate how acts of discrimination and hate crimes against the Asian community have impacted the mental health of young Asian Americans. This review seeks to explore the many effects of race-based discrimiation specifically for Asian Americans during and after the COVID-19 (coronavirus pandemic) such as: general feelings of inclusivity, physiological responses as a result of increasing racist encounters, and incidences of mental health experiences. Overall, the paper highlights how these characteristics manifest within the Asian American population and what measures and interventions can be done to protect Asian American from the negative consequences of hate.

## Introduction

COVID-19 is a global pandemic that has changed the lives of millions of people. Since the pandemic began, over 5 million people from over 200 countries around the world have been affected, causing both physiological and psychological determinants [1]. In particular, Asian Americans & Pacific Islanders (AAPI) have faced over 7,000 hate incidents, marking a 150% increase in 2020 and 194% increase in the first quarter of 2021, totaling 344% since 2019. Implicit bias and explicit anti-Asian sentiments have been fueled by references to the coronavirus as the “Chinese virus” or “Kung Flu” by government officials and other media, dramatically increasing anti-Asian racism and xenophobia worldwide [2]. As a result, the recent instances of hate crimes and discrimination during the COVID-19 pandemic may create unique, long lasting mental health challenges for the Asian American community. This can result in physiological comorbidities as data has shown how factors such as stress have led to as large as an 1.6-fold increase in risk of coronary heart disease and increased incidence in cancer. With studies having shown how symptoms such as anxiety, panic, paranoia, and depression have increased throughout the pandemic, this highlights cause for concern in lingering long term consequences for the future [3]. Extensive research into the mental health effects of this racism will provide further quantification of its impact on the AAPI population as well as methods for clinicians and health care systems to better care for the Asian American community affected by this discrimination [4].

## Methods

### Overview

To elucidate the relationships between discrimination and hate crimes throughout history on the mental health of adolescents within the Asian American population, a structured literature review was conducted with three databases employed to find literature pertaining to this subject: NCBI, Google Scholar, and PubMed.

### Search Term Strategy

Various search terms were used in this study for each database and article selected. The terms included: “Asian American and Pacific Islander” OR “AAPI” OR “Asian American” OR “Anti-American” OR “Anti-Asian” AND “COVID-19” OR “Pandemic” AND “youth” OR “young” AND “racial discrimination” OR “mental health” OR “PTSD” OR “cardiovascular health” OR “anxiety” OR “depression” OR “substance disorders” OR “physiological stress symptoms and behaviors” AND “inequality” OR “stress” OR “microaggressions” OR “hate crime” AND “model minority” OR “therapy” OR “counseling.” During the search process, additional terms were added or omitted as needed.

### Inclusion Criteria

For this research paper, studies were included that focused on meta-analysis, literature reviews, and primary research to investigate the associations between the mental health of young Asian Americans older than 16 and discrimination and hate crimes during the COVID-19 pandemic.

### Types of Studies

A variety of research papers were used for the purpose of this study including, but not limited to: primary review articles, meta-analyses, cohort studies, cross-sectional surveys, systematic reviews, and regression models.

### Types of Outcome Measures

The amount of people with mental health issues and experiences of discrimination and microaggressions was qualitatively assessed.

### Data Extraction

The time period of the papers used in this study ranged from 2011 to 2021. Information from the articles including the methods, data, results, and conclusions were extracted from each paper. The data included information about, sample size, demographics, and both qualitative and quantitative analysis for outcomes.

### Data Analysis

Data was extracted from each source and analyzed and extrapolated. The analysis of the data will be presented in the results and discussions section of the literature review.

## Results

### History of Hate Crime Towards Asian Americans In the United States

In our meta-analysis, the historical beginnings of Anti-Asian discrimination were first examined starting as early as the 1800s during the era of the Chinese Exclusion Act of 1829. During the 2003 SARS epidemic, 14% of Americans avoided Asian businesses and Asian Americans experienced increased threat and anxiety during SARS [5]. But this does not compare to the 31.7% of parents and 45.7% of youth who reported experiencing direct racial discrimination online and 50.9% of parents and 50.2% of youth in-person [6]. Data from Stop AAPI Hate further divides this in-person discrimination into verbal and physical with 70% being verbal. According to the Asian American Racism-Related Stress Inventory (AARRSI), conducted by Miller et al., racism was further divided into Socio-Historical, General, and Perpetual Foreigner Racism with scores of 0.82-0.93, 0.75-0.87, and 0.84-0.88, respectively, with higher scores indicative of higher levels of racism-related stress [7].

From the Riverside Acculturation Stress (RAS) Inventory which participants were asked to measure cultural stressors on a scale of of 1-5 (1 being strongly agree and 5 being strongly disagree), results include 0.82 for a RAS total score, 0.72 from the work challenge category, 0.75 for language skills, 0.71 for intercultural relations, and 0.70 for cultural isolation [7]. From the Bicultural self-efficacy (BSE) scale which is a 14-item self-report measure that assesses Asian American confidence with interactions and perceived incompatibility, results were 0.90 for BSE total score, 0.89 for nuances, 0.90 for bilingual skills and 0.87 for bicultural pressures subscales [7]. In the Mental Health Inventory, participants were assessed for both positive and negative psychological health outcomes. Items in the survey were rated on a 6 point scale, resulting in 0.91 for anxiety and 0.94 for depression scales [7]. From each of these diverse surverys and inventories, there seems to be a direct correlation between stress and other negative behavioral outcomes among Asian Americans and racism or other discriminatory behavior.

Furthermore, it was found that hate crimes could then be subdivided into different themes. These themes ranged from “ascription of intelligence,” “invalidation of interethnic differences,” and “Invisibility.” For example, “a degree of intelligence is assigned to an Asian American based on his/her race.” That example is one that fits under “ascription of intelligence” which would then “create tensions between her and other Black and Latino coworkers” [8]. An example under “invisibility” occurred when “Asian American individuals of all ethnic groups shared that they were often left out whenever issues of race were discussed or acknowledged” and that “Asian participants felt trapped in that when issues of race are discussed, they were considered like Whites, but never fully accepted by their White peers” [8]. It was also discussed that “hate crimes during the COVID-19 pandemic are an extreme manifestation of othering illustrating the replicative and cumulative effects of the historical embeddedness of racism and xenophobia. That is, perpetrators exact violence to dehumanize and ostracize Asian Americans, stigmatizing and “marking” them as deviants to destroy their sense of belonging” [4]. The amalgamation of these events lead to increased events of stress, post-traumatic stress, and a feeling of alienation in Asian Americans [9].

### Consequences of Hate Crime on the Mental Health of Asian Americans

Along with the presence of hate crime during the COVID-19 pandemic, there are significant consequences on the mental health of Asian Americans. There are concerns specifically about anxiety, substance abuse, depression, and stress among Asian Americans. In addition, it is important to highlight that there are general cultural factors that can influence Asian American mental health. Kramer et al. noted several cultural aspects among Asian Americans regarding mental health that may be in conflict with certain beliefs that are held in the United States. These different views along with traditional and natural methods for healing may be a possible source of discrimination as others, especially medical practitioners and providers, may not be aware or have a proper understanding of such beliefs [10]. Lee et al. interviewed different communities of Asian Americans and found a variety of common stressors that impacted mental health in Asian American young adults such as discrimination due to racial background, obligation to obtain high academics, balancing two cultures in America, and familial pressures [6]. Researchers found that Asian American young adults are less likely to seek mental health services; however, they are more likely to utilize personal support networks [6]. Since Asian cultures typically do not value the importance of mental health, this may play a factor in decreased help-seeking behavior [6].

#### Anxiety

From the study, Cheah et al, showed that “overall, 18.7 to 23.91% of the youth had a slightly elevated to substantial risk of clinically significant mental health problems, higher than the US norm.”[2]11.3% of youth in the sample reporting anxiety symptoms at moderate to severe levels, similar to that of Chinese youth during the COVID-19 outbreak in China (10.4%) [2]. From these results, it can be inferred that Asian American youth are more likely to develop mental health issues that arose from the discrimination during the COVID-19 pandemic era [2]. Rajita Sinha had studied the association between stressful events as a predictor of alcohol and drug dependence showing how stress was a key indicator in addiction vulnerability and cravings [11]. Specifically, they noted how adolescents are likely to also show decreased emotional, behavioral and self-control to increase risks of substance abuse and maladaptive behavior [11]. This can be a cause of concern in Asian American populations according to Fong et al. They had believed that AAPI populations may be less likely to ask for help for themselves in regards to substance abuse as nearly 40% of Asian callers to tobacco helplines and similar programs were from friends and family of affected individuals - compared to only 6% for non-AAPI populations [12]. Recent studies have also shown that AAPI substance dependent patients are much less likely to enter substance abuse treatment (8%) compared to non-AAPI patients (16%) [12].

#### Depression

From analyzing various studies, there is a strong correlation between acts of discrimination and hate crime with anxiety and depression among Asian Americans during the COVID-19 pandemic. It was found that self-reported vicarious racism and racial discrimination vigilance correlated with symptoms of depression and anxiety among Asian Americans. The article referred to vicarious racism as “witnessing or hearing about members of the same racial group experiencing racism during the COVID-19 pandemic,” and referred to racial discrimination vigilance as “efforts to prepare for, anticipate, or avoid being the target of racial discrimination during the COVID-19 pandemic” [1]. 91.9% of Asian participants in the study experienced vicarious racism in general, and 51.0% of Asian participants reported their racism experiences during the COVID-19 pandemic were more frequent than usual. 89.6% of Asian participants also reported being at least somewhat distressed from these experiences [1].

In addition, Paradies et al. studied research papers ranging from 1983 to 2013 and found that the highest reported mental health outcome associated with racism was depression. Depression was reported in 37.2% of articles, which was followed by psychological stress, which was reported in 21.3% of articles, and anxiety, which was reported in 14.4% of articles [13]. Dubey et al. conducted an analysis on the psychosocial impact on the COVID-19 pandemic. It discussed information about how the virus and nationwide lockdowns produced acute panic, anxiety, depression, and post-traumatic stress disorder (PTSD) [3]. These experiences have been fueled by xenophobia, racism, and stigmatization against communities who are “Chinese-looking” [3]. Specifically, these instances were frequently shared through social media platforms [3].

Lee et al. studied the interaction between anxiety, depressive Symptoms, physical symptoms and sleep difficulties within AAPI populations. The data revealed that 29% of participants reported increased discrimination, 41% reported increased anxiety, 53% had increased depressive symptoms, 15% had increased physical symptoms and 43% reported increased sleeping difficulties [14]. Overall, there has been a strong link among Covid-19 impact on depressive and anxiety symptoms within the AAPI population based on their regression models [14].

#### Stress

Using cross-sectional questionnaires, researchers found that Asian Americans experienced major mental health disparities and greater psychological distress compared to White Americans [15]. In addition, only 9.6% of Asian Americans reported prior use of mental health services, while 25.3% of White Americans reported prior use of mental health services [15].

Lee et al. reports the results of a study that measured levels of loneliness, depression, and anxiety among young adults. The results indicated that there was a slight increase from January to April/May in all three areas, yet that is still concerning as these negative behavioral outcomes are projected to peak even beyond this time frame. Results also indicate that women were more likely than men to experience loneliness [16]. The results also showed interesting outcomes in that young adults with stronger social networks felt greater disruptions to their networks and ultimately felt greater amounts of loneliness [16].

Wong-Padoongpatt et al. discussed the study that was performed on Asian American participants, with the first part interacting with a white perpetrator and the second conducting a survey about their levels of stress. Results indicated that self reported stress and physiological stress were not related to each other, and that Asian Americans felt higher levels of stress when interacting with a White American perpetrator compared to an Asian American perpetrator. Overall, these higher levels of stress were also associated with decreased levels of implicit self-esteem. These results suggest that the participants during the study tended to internalize the situation rather than saying the cause is due to external factors. This too, contributes to increased levels of physiological stress. Overall, results indicated that Asian Americans experience higher levels of stress and other negative behavioral outcomes when faced with a white perpetrator which indicates feelings of inferiority that results in these psychological and physiological changes that were recorded.

### Potential Physical Manifestations of Stressors in Adolescent Asian Americans

Along with the impact on mental health within the adolescent Asian American population, while not thoroughly studied yet, there is cause for concern regarding physical manifestations of chronic diseases in the long term. Kiviki et al. studied the impact of various stressors and their impact on the cardiovascular health of adults. After conducting a meta-analysis of large cohort studies, they found how cardiovascular disease within the general population has a moderate-strong association when having experienced extreme stress in one’s childhood [17]. They also found that adults with work or private-life stress have between a 1.1 to 1.6-fold increase in risk of coronary heart disease and stroke [17].

Along with increased risk for cardiovascular disease, stressors such as experiences in racism can lead to poorer health in terms of obesity. Paradies et al. conducted a meta-analysis with research articles from 1983 to 2013 which examined racism as a determinant of health. The study found that racism is significantly related to poorer health, with the strength of the association having a two-fold range [13]. Although the relationship is stronger for mental health than physical health, longitudinal studies found that physical health, overweight-related outcomes, such as Body Mass Index (BMI), obesity, overweight, waist circumference, and waist-hip ratio were significantly related to racism [13]. The study suggested that exposure to racist experiences may have led to hypothalamic-pituitary-adrenal (HPA) axis dysregulation that can damage the body and affect physical health such as cardiovascular disease and obesity [13].

Massetti et al. had also studied the impact of worsening mental health on health outcomes regarding diabetes and cancer. Young adult men and women aged 18-39 with mental health problems had worse health: they were more likely to report a history of asthma, skin cancer, diabetes, having ≥5 days in poor physical health in the past month, and having fair to poor health [18]. For diabetes in particular, 3.6% of men (95% CI: 1.03-1.17) with mental health issues had diagnosed diabetes compared to 1.4% of men (95% CI: 1.3-1.7) without. For women, it was 4.1% (95% CI: 3.5-4.7) with and 1.7% (95% CI: 1.4-1.9) without. Risk factors for cancer were also compared in both men and women with people suffering from mental health problems significantly more likely to smoke, binge drink, sleep inadequately or have no physical activity. After controlling for health status, demographic characteristics, and other confounding variables, only cervical cancer screening was not associated with mental health issues [18]. Overall, both indirect and direct forms of discrimination and hate crimes against the Asian community significantly impact the mental health of young Asian Americans. Our results uncovered more information and insight on this important issue and its effect on the mental health of these populations.

## Discussion

History plays an important role in defining the behaviors and viewpoints of Asian American today. Discrimination towards Asian Americans was noted as early as the 1800s during the era of the Chinese Exclusion Act and continued on during the 2000s during the SARS Epidemic. During these times, not only were individual people affected but also businesses and any Asian establishments experienced increased threat and anxiety. But the COVID-19 pandemic brought this to a higher level with over 45% and 50% of youth experiencing direct racial discrimination online and in-person, respectively with in-person further divided into 70% verbal and 30% physical.

Along with the presence of hate crime during the COVID-19 pandemic, there are significant consequences on the mental health of Asian Americans. There are concerns specifically about anxiety, substance abuse, depression, and stress among Asian Americans. Asian American youth exposed to discrimination demonstrated slightly elevated to substantial risk of clinically significant mental health problems, higher than the US norm. 11.3% of youth in the sample reporting anxiety symptoms at moderate to severe levels, similar to that of Chinese youth during the COVID-19 outbreak in China (10.4%). From these results, it can be inferred that Asian American youth are more likely to develop mental health issues that arose from the discrimination during the COVID-19 pandemic era.

The metaanalysis found that the discrimination experienced by Asian American’s throughout history can impact their mental health in various ways. From discrimination experienced through the COVID-19 Pandemic itself, Cheah et. al showed that 11.3% of Asian American youth in the United States reported anxiety symptoms at moderate to severe levels, with as much as 23.91% of Asian American youth being at elevated risk for significant mental health ailments overall [2]. These studies had also found an increased incidence in depression within this population as a result of acts of discrimination. A rapid increase in self reports of racism/racial discrimination correlated with symptoms of depression felt among Asian American’s with many reporting distress from these events. Paradies et. al had found that depression was the highest reported mental health outcome associated with racism, much of which was fueled through social media platforms and acts of violence all around the country against the Asian American population [3,18]. Lee et. al even reported that AAPI populations had shown an 53% increase in depressive symptoms alongside anxiety when experiencing discrimination [14]. This is of concern as increased anxiety and depression can lead to maladaptive behaviors such as substance abuse, where Sinha et. al had shown that there was a strong association between stressful events with alcohol and drug dependency within adolescents [11].

Overall, this is cause for concern long term as these mental health manifestations can lead to long term consequences due to poor coping mechanisms. Overall Asian Amercans have been found to show greater psychological distress compared to white americans. This increase in stress has been linked to decreased levels of implicit self esteem, causing an increased incidence in stress levels at any given moment which can lead to impaired mood, social reclusion, impact on performance and much more. This is only made worse by the fact that only 9.6% of Asian Americans had reported prior use of mental health services, something that Sinha et. al alluded to with AAPI populations being less likely to look for help for their problems [15,20]. Overall, this increase in anxiety, depression, substance abuse and stress puts Asian American adolescents at risk for adverse outcomes in the future.

Alongside worsening mental health, stressors in Asian Americans also manifested physical symptoms. Asian Americans with elevated stress due to a history of hate crimes were more likely to report symptoms of asthma, skin cancer, diabetes; furthermore, 3.6% of men and 4.1% of women with mental health problems had diagnosed diabetes compared to 1.4% of men and 1.7% women without reported mental health problems [15]. As evident by these results, elevated stress can lead to serious chronic health conditions. Stress also manifested other associated conditions such as smoking, binge drinking, inadequate sleep, and obesity that can further worsen an individual’s health [13]. A full regression model also found that COVID-19 impact on Asian American’s physical status was statistically significant, meaning that the discrimination incurred due to COVID-19 had a direct measurable impact on their physical health [14]. Decreased mental health, coupled with increased stress lead to numerous physical manifestations that can lead an individual down a dangerous spiral downwards with one factor (example: being diagnosed with a chronic condition) leading to another.

COVID-19 has had a large effect on the mental and physical health of the general population. With the already present difficulty, rising hate crimes and discrimination towards Asian Americans worsened and further complicated their mental and physical health. This presented itself in a myriad of ways, primarily by utilizing other unhealthy coping methods - further worsening their physical and psychological health. By targeting rising hate crimes/discriminatory actions, Asian Americans can find reprieve from this source of stress which will directly positively affect their physical health.

## Limitations

Although a wide range of papers were examined for this study, including research using primary review articles, meta-analyses, cohort studies, cross-sectional surveys, and systematic reviews, there are limitations to this study. Generalizing the results of these studies is currently limited. The impact on physical health specifically within the young Asian American population is not thoroughly studied yet. For example, while there are studies that indicate a general association between discrimination and physical manifestations, the effect in specifically young Asian Americans has not been extensively researched and requires further examination. Upon literature search, the relationship between discrimination and other chronic diseases in Asian Americans, including diabetes, cardiovascular disease, and cancer, did not appear in current literature.

## Conclusion and Implications

To improve the mental health of young Asian Americans especially during the post-COVID, there needs to be attention given to both the triggers and the physiological manifestations. During the height of COVID-19 there were multiple identifiable triggers that affected the mental health of young Asian Americans. Instances of hate crime are main causes and studies have shown that when encountered by their white counterparts, Asian Americans felt increasing amounts of stress and other negative psychological events, coupled with feelings of inferiority. It is evident that negative psychological and physiological outcomes are concurrent, with Asian Americans reporting increased levels of stress, anxiety and depression that can also potentially develop into cardiovascular and metabolic manifestations. Behavioral problems can also arise as a result of discriminatory behavior that can alter self perception. Overall, the COVID-19 pandemic has highlighted and even increased the negative outcomes and manifestations that can arise from acts of racism and discrimination.

The implications of the results discussed and analyzed identify that Asian Americans experience more amounts of stress due to discriminatory events and this has heightened due in the COVID-19 pandemic area. Therefore, there needs to be more attention to Asian American mental health and regular wellness visits such as in the primary care setting. Following this, there needs to be attention from physicians and other healthcare workers to be familiar with certain triggers and offer support and resources to young Asain Americans and especially if they are not seeking it themselves.

## Data Availability

No data gathered in this study.

## Acknowledgements

None

